# ROS1 immunohistochemistry as a potential predictive biomarker for ROS1-targeted therapy in breast cancer: impact of antibody clone selection

**DOI:** 10.1101/2024.12.09.24318695

**Authors:** Anna Sokolova, Vaibhavi Joshi, Haarika Chittoory, Michael Walsh, Malcolm Lim, Jamie R. Kutasovic, Kaltin Ferguson, Peter T. Simpson, Sunil R. Lakhani, Amy E. McCart Reed

## Abstract

**Aims:** Invasive lobular carcinoma (ILC) may show targetable vulnerabilities secondary to the characteristic loss of the cell adhesion protein E-cadherin. Specifically, a synthetic lethal interaction was identified between E-cadherin loss and ROS1 inhibition. Several clinical trials are currently underway to assess the efficacy of ROS1 inhibitors in ILC, however, ROS1 expression has not been confirmed in ILC tumours and ROS1 has not been validated as a biomarker in the breast cancer setting. This study aimed to (i) examine ROS1 expression in a large cohort of breast cancer cases and (ii) investigate the biology and clinical significance of ROS1 positivity in breast cancer.

**Methods:** ROS1 immunohistochemistry was performed on a large cohort of ILC (n=274) and invasive carcinoma of no special type (NST; n=431) cases with extensive clinicopathological data. The staining performance of four ROS1 antibody clones was compared.

**Results:** There was marked variation in ROS1 status according to antibody clone. D4D6 and SP384 were negative in almost all breast cancer cases, whereas EP282 and EPMGHR2 were positive in 37% and 47% of ILC cases, and 49% and 74% of NST cases, respectively. Only data from clones D4D6 and SP384 were highly concordant, while EP282 and EPMGHR2 were positive in distinct breast cancer subtypes.

**Conclusions:** Assessment of ROS1 status in breast cancer appears to be highly antibody clone dependent. ROS1 antibody clone selection will be an important consideration in the design of clinical trials investigating the clinical validity of ROS1 as a predictive biomarker in breast cancer.

## Introduction

Breast cancer is the most common malignancy globally.^1,2^ The predominant histologic subtype is invasive carcinoma of no special type (NST), which comprises up to 75% of all breast cancer cases.^3^ Invasive lobular carcinoma (ILC) is the second most common subtype, constituting up to a further 15% of cases.^3,4^ ILC and NST demonstrate distinct clinicopathological and molecular features and differ in their therapeutic response and prognostic outcomes.^5^ Despite these clinical and biological differences, there is a lack of treatment approaches specifically tailored to ILC patients, with an unmet need for translational research that focuses on the optimisation of therapeutic strategies for this patient subgroup.^6^ Recently, synthetic lethality has emerged as a potentially effective targeted treatment strategy for ILC tumours.^7–9^ This treatment approach exploits the relationship between two interdependent genes, where simultaneous disruption causes cell death.^10,11^ The loss of the cell adhesion protein E-cadherin is a defining feature of lobular neoplasms^12,13^ and is associated with synthetic lethal vulnerabilities in ILC cells.^7,8^ Indeed, a particularly robust synthetic lethal interaction has been identified between E-cadherin loss and the inhibition of the receptor tyrosine kinase ROS1, resulting in targeted tumour cell death in multiple E-cadherin negative breast cancer models.^9^

ROS1, encoded by the proto-oncogene ROS1 located on chromosome 6, is part of the insulin receptor family of receptor tyrosine kinases (RTKs).^14,15^ In humans, ROS1 is predominantly expressed in the lung, with lower gene expression in the brain, kidney and testis.^16^ ROS1 is considered an orphan RTK, because its endogenous activating ligand has not been definitively characterised. Functional studies of oncogenic ROS1 fusion proteins have shown that aberrant ROS1 activation is associated with signalling of several downstream oncogenic pathways involved in cell survival, growth and proliferation.^17–21^ ROS1 fusion genes are dominant oncogenic drivers in 1-2% of non-small cell lung cancers (NSCLC),^22–24^ however are rarely seen in breast cancer (in up to 0.04% of cases).^25,26^ While other ROS1 alterations such as copy number and nucleotide variations are found across different tumour types, their oncogenic potential is unclear.^22^ Accordingly, ROS1 targeted therapy is only licensed for ROS1-rearranged lung cancers at present, however the novel synthetic lethality data suggests that ILC and other E-cadherin negative breast cancers may also benefit from ROS1 inhibitor therapy. This is based on interdependence between ROS1 and E-cadherin in regulating cytokinesis, with ROS1 inhibition leading to mitotic catastrophe and cell death in E-cadherin deficient cells.^9^

Three phase II clinical trials are currently exploring the potential role of various ROS1 inhibitors, combined with endocrine therapy, in advanced/metastatic ILC (ROLo, NCT03620643 and REPLOT, NCT06408168) and early ILC (ROSALINE, NCT04551495). Confirmation of a robust predictive biomarker for E-cadherin/ROS1 synthetic lethality is needed to guide patient management to identify potential responders and eliminate overtreatment.^27^ However, there is currently no validated predictive biomarker for E-cadherin/ROS1 synthetic lethality. Indeed, the drug target ROS1 has not been well characterised in breast cancer cases and ROS1 expression has not been confirmed in ILC. Ostensibly, characterising ROS1 protein expression in E-cadherin negative breast cancers is central to identifying a patient cohort that is likely to respond to ROS1 inhibitor therapy based on synthetic lethality. Herein we present an assessment of ROS1 expression in a large cohort of ILC and NST cases comparing four different ROS1 biomarker candidates.

## Materials and Methods

### Ethics

Human Research Ethics Committees at The University of Queensland (2005000785), Royal Brisbane and Women’s Hospital (RBWH 2005/022) and the UnitingCare Health (2006-39) and Public Health Act from the Queensland Government (CT_4352) approved the study.

### Clinical samples

Formalin-fixed paraffin-embedded (FFPE) tumour blocks from Royal Brisbane and Women’s Hospital, Princess Alexandra Hospital and Wesley Hospital archives (Queensland, Australia); 0.6 µm cores were sampled in duplicate as tissue microarrays (TMAs)^28,29^ including 274 ILC and 431 NST cases with comprehensive clinicopathological annotation (Table 1).

**Table 1.** Summary of patient characteristics within the ILC and NST cohorts.

| Variable |  | ILC total<br>n (%) | NST total<br>n (%) |
| --- | --- | --- | --- |
| Age | <45 | 27 (10) | 74 (18) |
|  | 45-55 | 67 (25) | 106 (26) |
|  | >55 | 176 (65) | 231 (56) |
|  | Mean age | 61 years | 59 years |
| Size | T1 | 97 (43) | 135 (48) |
|  | T2 | 104 (46) | 120 (42) |
|  | T3 | 25 (11) | 28 (10) |
| Grade | 1 | 16 (6) | 59 (14) |
|  | 2 | 228 (84) | 167 (39) |
|  | 3 | 29 (11) | 204 (47) |
| LN status | Positive | 74 (42) | 86 (42) |
|  | Negative | 102 (58) | 117 (58) |
| ER | Positive | 255 (97) | 266 (63) |
|  | Negative | 8 (3) | 159 (37) |
| PR | Positive | 209 (84) | 225 (53) |
|  | Negative | 41 (16) | 200 (47) |
| HER2 | Positive | 6 (3) | 54 (13) |
|  | Negative | 213 (97) | 372 (87) |

### Immunohistochemistry

Table 2 shows ROS1 antibody and staining details. Immunohistochemistry with ROS1 antibody clones EP282 and EPMGHR2 was performed manually in-house, while SP384 and D4D6 clone staining was performed at an accredited diagnostic laboratory. Staining was visualized using the MACH1 HRP-Polymer kit (Biocare Medical, Concord, CA, USA). Positive controls included lung adenocarcinoma with confirmed ROS1-rearrangement and a commercially available ROS1 analyte control slides (HistoCyte, Abacus Dx, HCL023), comprising a positive (lung adenocarcinoma with a SLC34A2::ROS1 translocation) and a negative control core (ROS1 negative breast cancer cell line). ROS1 staining was scored according to the highest intensity (0=no staining, 1=weak staining, 2=moderate staining and 3=strong staining), with cytoplasmic and/or membranous staining patterns in ³5% of tumour cells considered positive. Cores with fewer than 10 unequivocal invasive tumour cells were excluded.

**Table 2.** Summary of ROS1 antibody clones and antigen retrieval conditions.

| Clone | D4D6 | SP384 | EP282 | EPMGHR2 |
| --- | --- | --- | --- | --- |
| Vendor | Cell signaling technology | Ventana | BioSB | Abcam |
| Host species | Rabbit monoclonal | Rabbit monoclonal | Rabbit monoclonal | Rabbit monoclonal |
| Format | RTU | RTU | Conc | Conc |
| Dilution | N/A | N/A | 1:100 | 1:300 |
| Platform | Automated | Automated | Manual | Manual |
| Epitope retrieval | Heat induced | Heat induced | Heat induced | Heat induced |
| Buffer | CC1 | CC1 | EDTA | EDTA |
| Incubation | 16min 36°C | 16min 36°C | Overnight 4°C | Overnight 4°C |
| Visualisation system | OptiView | OptiView | MACH1 | MACH1 |

RTU, ready-to-use; Conc., concentrated; CC1, cell conditioning 1 buffer (a ready-made Tris-EDTA solution).

**Table 3.** Summary of ROS1 staining in ILC cohort according to antibody clone.

| Cohort | Clone | Total cases | Staining intensity |  |  |  | Total positive (%) |
| --- | --- | --- | --- | --- | --- | --- | --- |
|  |  |  | Negative | 1+ | 2+ | 3+ |  |
| ILC | SP384 | 195 | 188<br>(96%) | 7<br>(4%) | 0 | 0 | 7<br>(4%) |
|  | D4D6 | 210 | 205<br>(98%) | 5<br>(2%) | 0 | 0 | 5<br>(2%) |
|  | EP282 | 203 | 108<br>(53%) | 53<br>(26%) | 32<br>(16%) | 10<br>(5%) | 95<br>(47%) |
|  | EPMGHR2 | 204 | 128<br>(63%) | 45<br>(22%) | 24<br>(11%) | 7<br>(3%) | 76<br>(37%) |
| NST | SP384 | 297 | 292<br>(98%) | 5<br>(2%) | 0 | 0 | 5<br>(2%) |
|  | EP282 | 293 | 149<br>(51%) | 78<br>(27%) | 39<br>(13%) | 27<br>(9%) | 144<br>(49%) |
|  | EPMGHR2 | 338 | 87<br>(26%) | 98<br>(29%) | 95<br>(28%) | 58<br>(17%) | 251<br>(74%) |

### Western Blot

HEK293T and HeLa cells (American Type Culture Collection) were transfected with 500 ng pcDNA3.1-ROS1-V5/HIS plasmid (Addgene; a gift from Pavel Krejčí ^30^) using Lipofectamine™ 3000 (Invitrogen). Control wells received transfection conditions without plasmid. Cells were resuspended in 1X RIPA buffer supplemented with 1% protease inhibitor. 20 µg of protein was resolved on a 4-12% bis-tris SDA-PAGE gel (Bolt, Invitrogen, ThermoScientific) and transferred to PVDF membrane (Millipore, MA, USA). After blocking with 5% BSA in Tris-buffered saline with 0.1% Tween-20 (Sigma Life Science), membranes were probed overnight with ROS1-D4D6 (Cell signalling technology; 1:1000), ROS1-EPMGHR2 (Abcam; 1:1000), ROS1-EP282 (BiosSB; 1:1000), and GAPDH (14C10, Cell signalling technology; 1:1000) at 4°C. Secondary antibody was goat anti-rabbit IgG FC-specific HRP (Sigma Aldrich; 1:20,000) and detection performed with SuperSignal ECL HRP substrate (West PicoPlus, ThermoScientific), and imaged with the Chemidoc MP imaging system (Biorad).

### Statistical Analyses

Statistical analyses were performed using GraphPad Prism (version 10.0, GraphPad Software) and AgreeStat360 (https://agreestat360.com). Relationships between ROS1 expression and clinicopathological parameters were assessed using chi-squared or Fischer’s exact tests, depending on variables number. Inter-assay reliability was determined by calculating percent observed agreement (total concordant cases expressed as a percentage of total cases) and Gwet’s agreement coefficient (AC1) which measures inter-rater reliability adjusting for chance in imbalanced data sets. Statistical significance was defined as p<0.05.

## Results

### ROS1 status and biological significance of ROS1 positivity according to antibody clone

ILC cases were stained with four ROS1 antibody clones (D4D6, SP384, EP282 and EPMGHR2) and NST cases were stained with three ROS1 antibody clones (SP384, EP282 and EPMGHR2) (Table 4). Examples of staining intensity are shown in Fig 1A; the results were highly variable across the antibody clones in both the ILC (Fig 1B) and NST cohorts (Fig 1C). The domain structure of ROS1, with the reported antibody binding sites is shown in Fig 1D. Clones SP384 and D4D6 were broadly negative in ILC. SP384 was similarly negative in NST, D4D6 was not tested in NST cases to preserve tissue. Clone EP282 was positive in 47% of ILC cases and 49% of NST cases, while EPMGHR2 was positive in 37% of ILC cases and 74% of NST cases.

**Fig 1.**
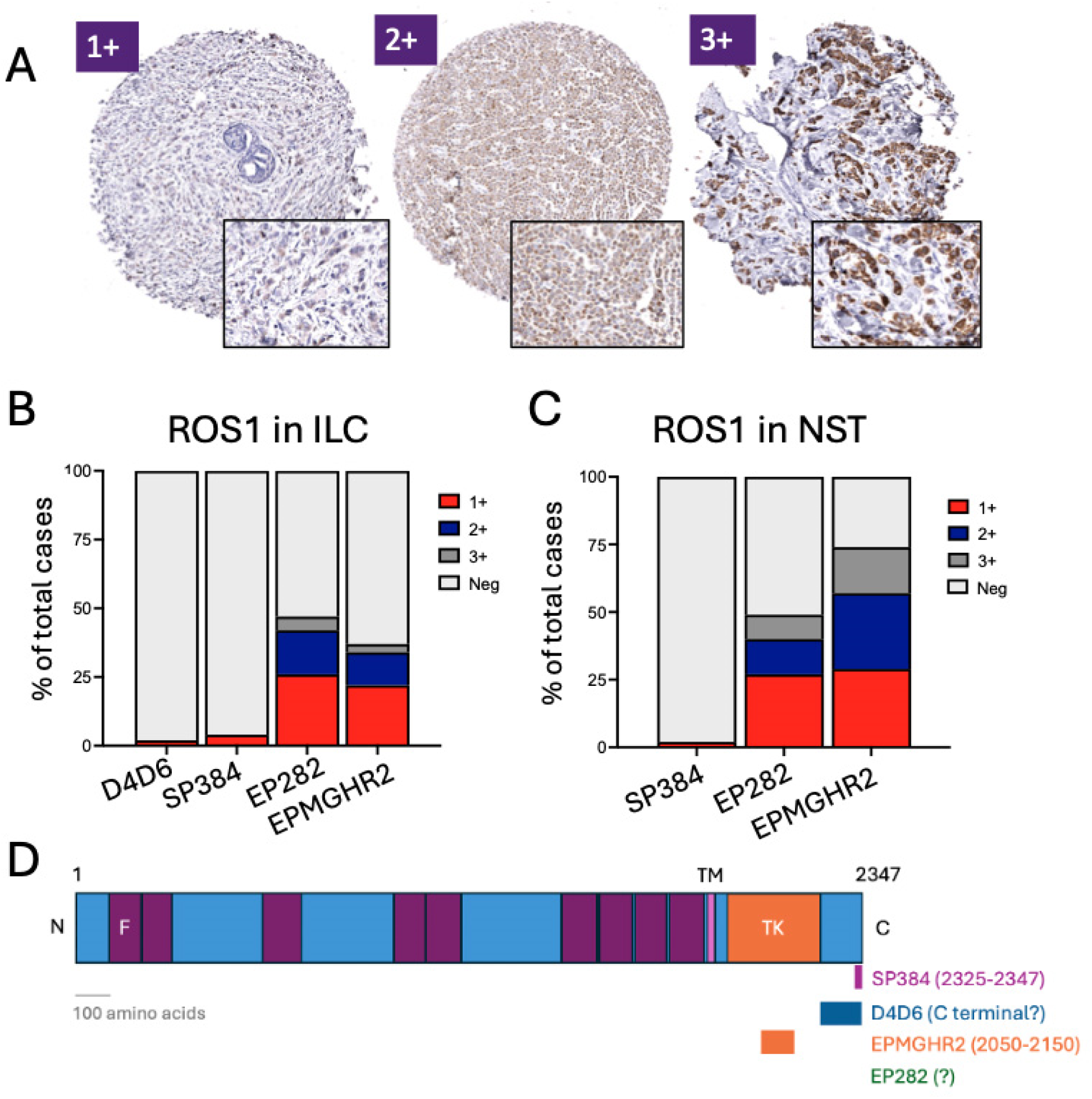
ROS1 expression as assessed by different antibody clones. Examples of staining intensity shown in (A), and the breakdown by clone in ILC (B) and NST (C). (D) Wild type ROS1 is a transmembrane protein with an extracellular N-terminal domain and an intracellular tyrosine kinase and C-terminal domain. Epitope binding sites of ROS1 antibody clones according to manufacturers are pictured (D4D6, Cell Signalling: C-terminal domain; SP384, Ventana: AA2325-2347, EPMGHR2, Abcam: AA2050-2150, EP282, BioSB: information on target region not available). N, N terminus; C, C terminus; TM, transmembrane; F, fibronectin type III; TK, tyrosine kinase

ROS1 expression detected with SP384 and D4D6 was not significantly associated with any clinicopathological parameters (Table 4, 5). While EP282 staining was not significantly associated with clinicopathological parameters in ILC (Table 4), there was a significant association with ER (p=0.0003) and PR (p=0.0018) negativity, HER2 positivity (p<0.0001) and higher tumour grade (p=0.0015) in NST (Table 5). EPMGHR2 staining showed a significant association with younger age (p=0.0307) in ILC (Table 4) while in NST, EPMGHR2 staining was significantly associated with ER (p=0.0091) and PR (p=0.0025) negativity, triple negative status (p=0.0131), and higher tumour grade (p=0.0001) (Table 5).

**Table 4.** Relationships between ROS1 positivity and clinicopathological variables according to antibody clone in ILC.

| Variable |  | D4D6 |  |  | SP384 |  |  | EP282 |  |  | EPMGHR2 |  |  |
| --- | --- | --- | --- | --- | --- | --- | --- | --- | --- | --- | --- | --- | --- |
|  |  | Pos | Neg | *P* | Pos | Neg | *P* | Pos | Neg | *P* | Pos | Neg | *P* |
| **Age** | <45 | 0 | 24 | 0.2193 | 0 | 24 | 0.4252 | 13 | 10 | 0.558 | 14 | 9 | **0.0307** |
|  | 45-55 | 5 | 125 |  | 6 | 118 |  | 57 | 71 |  | 41 | 86 |  |
|  | >55 | 0 | 53 |  | 1 | 45 |  | 24 | 26 |  | 20 | 30 |  |
| **Grade** | 1 | 0 | 10 | 0.7269 | 0 | 8 | 0.6997 | 2 | 5 | 0.1181 | 2 | 6 | 0.7420 |
|  | 2 | 4 | 173 |  | 5 | 59 |  | 78 | 95 |  | 64 | 107 |  |
|  | 3 | 1 | 22 |  | 2 | 21 |  | 15 | 8 |  | 10 | 15 |  |
| **Size** | T1 | 1 | 74 | 0.0567 | 4 | 66 | 0.3991 | 36 | 33 | 0.3462 | 26 | 47 | 0.3879 |
|  | T2 | 1 | 81 |  | 2 | 77 |  | 40 | 40 |  | 30 | 50 |  |
|  | T3 | 2 | 19 |  | 0 | 18 |  | 13 | 6 |  | 10 | 9 |  |
| **LN** | Pos | 1 | 56 | >0.9999 | 0 | 53 | 0.2583 | 33 | 25 | 0.8605 | 23 | 35 | 0.4801 |
|  | Neg | 1 | 75 |  | 3 | 67 |  | 40 | 33 |  | 34 | 39 |  |
| **ER** | Pos | 5 | 193 | >0.9999 | 7 | 174 | >0.9999 | 89 | 102 | 0.424 | 71 | 121 | 0.1083 |
|  | Neg | 0 | 7 |  | 0 | 7 |  | 4 | 2 |  | 5 | 2 |  |
| **PR** | Pos | 5 | 161 | >0.9999 | 4 | 147 | 0.2598 | 73 | 85 | >0.9999 | 57 | 103 | 0.2373 |
|  | Neg | 0 | 33 |  | 2 | 28 |  | 14 | 17 |  | 15 | 17 |  |
| **HER2** | Pos | 0 | 6 | >0.9999 | 0 | 6 | >0.9999 | 3 | 3 | >0.9999 | 1 | 5 | 0.4019 |
|  | Neg | 3 | 157 |  | 6 | 137 |  | 70 | 85 |  | 62 | 87 |  |

**Table 5.** Relationships between ROS1 positivity and clinicopathological variables according to antibody clone in NST.

| Variable |  | SP384 |  |  | EP282 |  |  | EPMGHR2 |  |  |
| --- | --- | --- | --- | --- | --- | --- | --- | --- | --- | --- |
|  |  | Pos | Neg | *P* | Pos | Neg | *P* | Pos | Neg | *P* |
| **Age** | <45 | 0 | 53 | 0.5926 | 24 | 28 | 0.9068 | 47 | 14 | 0.5138 |
|  | 45-55 | 2 | 69 |  | 34 | 36 |  | 55 | 24 |  |
|  | >55 | 3 | 156 |  | 78 | 79 |  | 137 | 44 |  |
| **Grade** | 1 | 1 | 38 | 0.3525 | 11 | 28 | **0.0015** | 25 | 17 | **0.0001** |
|  | 2 | 3 | 117 |  | 54 | 67 |  | 85 | 44 |  |
|  | 3 | 1 | 136 |  | 78 | 54 |  | 140 | 26 |  |
| **Size** | T1 | 0 | 87 | >0.9999 | 39 | 46 | 0.6181 | 71 | 28 | 0.7676 |
|  | T2 | 0 | 77 |  | 38 | 35 |  | 73 | 23 |  |
|  | T3 | 0 | 15 |  | 8 | 6 |  | 16 | 5 |  |
| **LN** | Pos | 0 | 53 | >0.9999 | 31 | 23 | 0.5761 | 58 | 12 | 0.1283 |
|  | Neg | 1 | 64 |  | 31 | 29 |  | 60 | 23 |  |
| **ER** | Pos | 4 | 206 | >0.9999 | 88 | 117 | **0.0003** | 149 | 65 | **0.0091** |
|  | Neg | 1 | 84 |  | 56 | 28 |  | 101 | 21 |  |
| **PR** | Pos | 3 | 173 | >0.9999 | 73 | 100 | **0.0018** | 123 | 58 | **0.0025** |
|  | Neg | 2 | 116 |  | 71 | 45 |  | 127 | 27 |  |
| **HER2** | Pos | 1 | 38 | 0.5117 | 32 | 8 | **<0.0001** | 30 | 13 | 0.4545 |
|  | Neg | 4 | 251 |  | 111 | 139 |  | 220 | 72 |  |
| **TNBC** | Yes | 0 | 61 | 0.5873 | 34 | 25 | 0.1902 | 82 | 16 | **0.0131** |
|  | No | 5 | 228 |  | 109 | 120 |  | 167 | 70 |  |

### Concordance between ROS1 antibodies

A single case (<0.1%) was classified as positive with all four clones in ILC (n=166; Fig 2) and all three clones in NST (concordant positive; n=260; Fig 3). In ILC, 67 cases (40%) were negative with all four clones and in NST, 55 cases (21%) were negative with all three clones (concordant negative). While 98 ILC (60%) and 204 NST (78%) showed discordant positive and negative staining. The overall observed agreement, including all concordant positive and negative results was 40% in ILC and 21% in NST.

**Fig 2.**
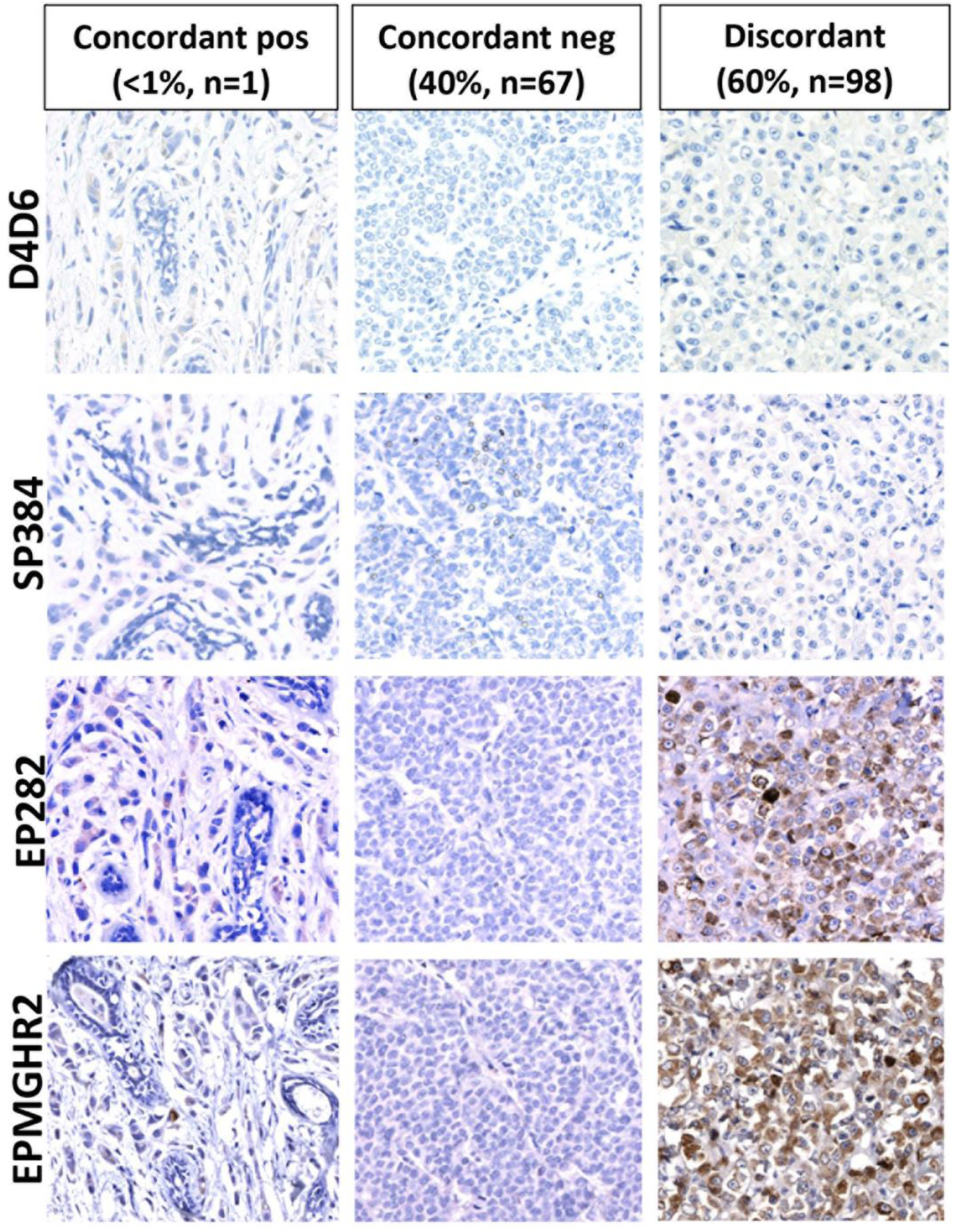
Examples of concordant and discordant ROS1 staining in ILC (total cases n=166).

**Fig 3.**
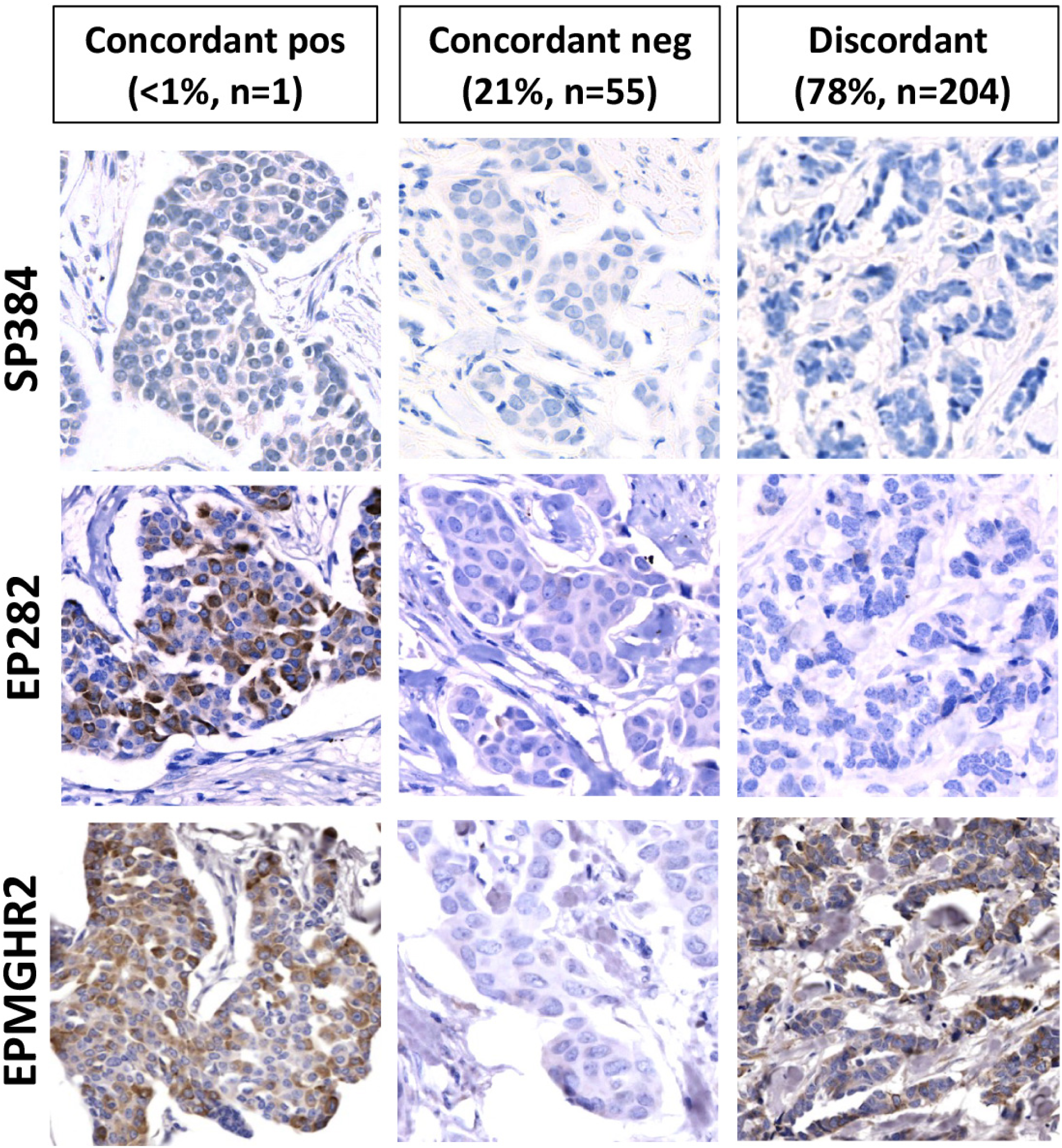
Examples of concordant and discordant ROS1 staining in NST (total cases n=260).

Comparing antibody pairs in ILC (Table 6), clones D4D6 and SP384 were highly concordant (agreement 96%, AC1=0.955). Of the negative cases (n=165), D4D6 and SP384 were both negative in 95.8% of cases (Fig 4A). Of the 8 cases classified as positive by both antibodies, there was concordant positive staining in one case and discordant staining in 7 cases (Fig 4B). In contrast, EP282 and EPMGHR2 were only moderately concordant (agreement 75%, AC1=0.497) (Table 6). While these antibodies were positive in a large proportion of cases, they were not consistently positive in the same cases. Of the positive cases (n=98), the two antibody clones were concordant in 57.1%; while in the negative cases (n=110), the two antibody clones were concordant in 61.8% of cases (Fig 4D,E). The other assay comparisons showed lower levels of agreement (Table 6).

**Fig. 4.**
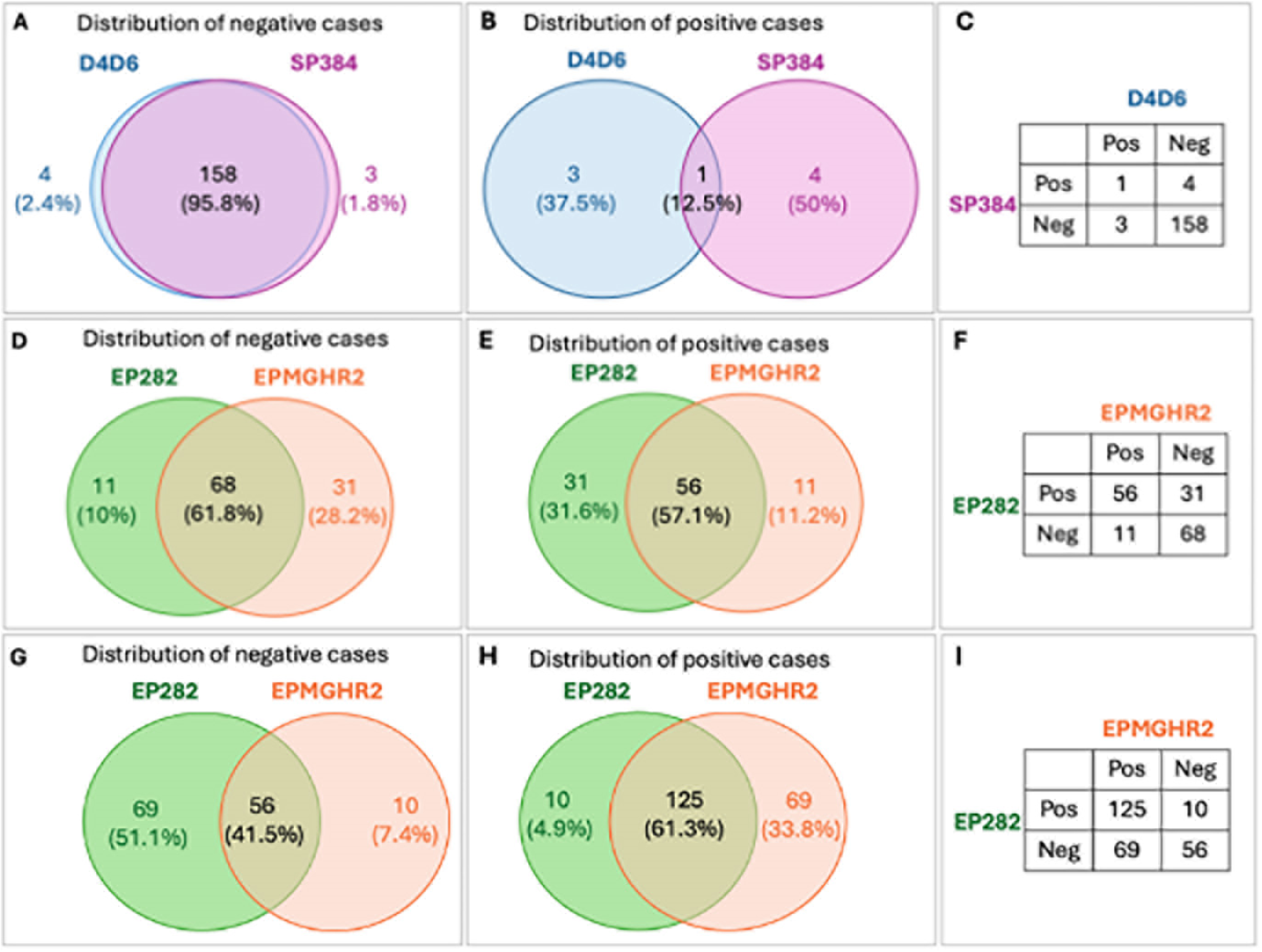
Comparisons of agreement in the two most concordant antibody pairs in ILC. **A**. Area-proportional Venn diagram depicting very high agreement for negative cases with D4D6 and SP384 (n=165). **B**. Area-proportional Venn diagram depicting the distribution of positive cases with D4D6 and SP384 (n=8). **C**. Confusion matrix comparing positive and negative results with D4D6 and SP384. **D**. Area-proportional Venn diagram depicting moderate agreement for negative cases with EP282 and EPMGHR2 (n=98). **E**. Area-proportional Venn diagram depicting moderate agreement for positive cases with EP282 and EPMGHR2 (n=110). **F**. Confusion matrix comparing positive and negative results with EP282 and EPMGHR2. Comparisons of agreement in the most concordant antibody pair in NST. **G**. Area-proportional Venn diagram depicting moderate agreement for negative cases with EP282 and EPMGHR2 (n=98). **H** Area-proportional Venn diagram depicting moderate agreement for positive cases with EP282 and EPMGHR2 (n=110). **I.** Confusion matrix comparing positive and negative results with EP282 and EPMGHR2.

**Table 6.** Level of agreement between antibody pairs (observed agreement %, Gwet’s AC1).

| Clone | Invasive lobular Carcinoma |  |  |  | IC-No Special Type |  |  |
| --- | --- | --- | --- | --- | --- | --- | --- |
|  | D4D6 | SP384 | EP282 | EPMGHR2 | SP384 | EP282 | EPMGHR2 |
| **D4D6** | 1.000 |  |  |  |  |  |  |
| **SP384** | 96%<br>0.955 | 1.000 |  |  | 1.000 |  |  |
| **EP282** | 49%<br>0.150 | 48%<br>0.136 | 1.000 |  | 47%<br>0.134 | 1.000 |  |
| **EPMGHR2** | 58%<br>0.374 | 60%<br>0.398 | 75%<br>0.497 | 1.000 | 26%<br>-0.397 | 70%<br>0.432 | 1.000 |

Of the three antibodies tested in NST (SP384, EP282 and EPMGHR2), the highest level of agreement was between clones EP282 and EPMGHR2, however similarly to ILC, these antibodies were positive in different patient subsets and only moderately concordant (agreement 70%, AC1=0.432) (Table6). Of the positive cases (n=204), these two clones were concordant in 61.3% and of the negative cases (n=135), these two clones were concordant in 41.5% of cases (Fig 4G,H). The other assay comparisons showed lower levels of agreement (Table6).

### Antibody Specificity

We assessed antibody specificity by overexpressing ROS1 in mammalian cell lines and performing a western blot (Fig 5). While D4D6 identified multiple bands in HEK293T, a single ∼260 KDa band was resolved in HeLa cells. EP282 and EPMGHR2 both detected a ∼260 KDa band in HEK293T but not HeLa cells. SP384 was unable to be tested as it is an automated Ventana diagnostic formulation.

**Figure 5.**
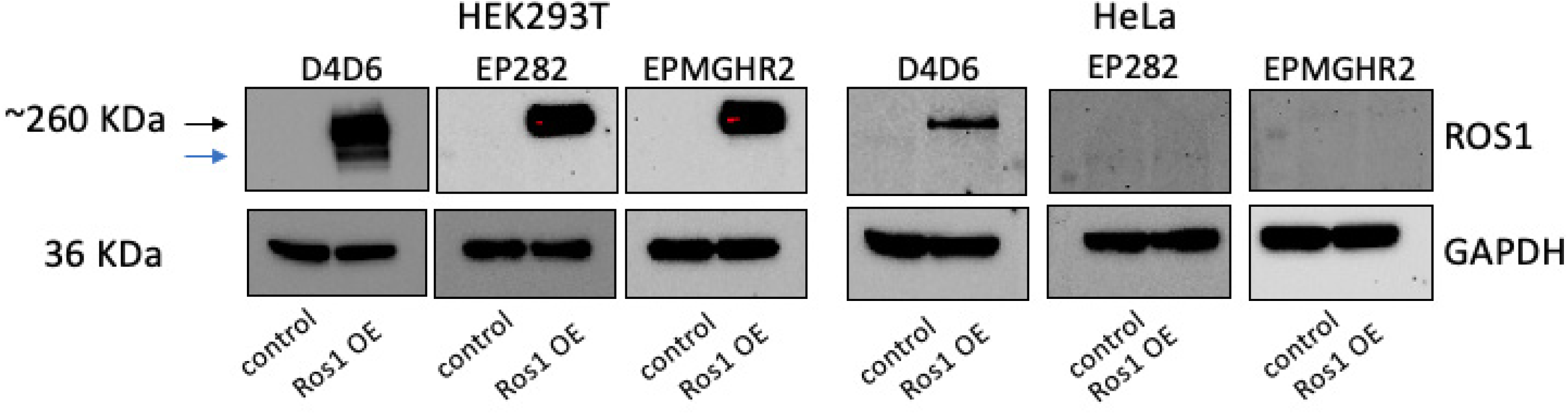
Specificity of ROS1 antibodies. ROS1 was overexpressed (OE) in HEK293T and HeLa cells, and protein expression studied via western blot. Black arrow identifies ROAS1 band, while the blue arrow highlights the non-specific band as detected by D4D6.

## Discussion

The efficacy of ROS1 targeted therapy in ILC is under evaluation in clinical trials, however ROS1 has not yet been validated as a biomarker in the breast cancer setting. To this end, we compared the performance of four different ROS1 antibody clones across a large cohort of breast cancer cases and assessed the potential biological significance of ROS1 positivity in breast cancer.

The performance of D4D6 and SP384 clones is clinically relevant as both are widely used in clinical practice to screen for ROS1 rearrangements in NSCLCs.^31^ The sensitivity and specificity of these clones has been derived from comparisons with gold standard molecular tests used to diagnose ROS1 rearrangements. We show that both D4D6 and SP384 were negative in almost all (>95%) breast cancer cases, with very few cases showing weak staining of uncertain clinical significance. The results obtained by these two antibodies were highly concordant, with 96% observed agreement and an agreement coefficient (AC1) of 0.955 indicating almost perfect agreement. Interestingly, both clones target the intracellular portion of the ROS1 protein at the C-terminal domain with likely overlapping epitopes (Fig 1), however the exact amino acid region targeted by D4D6 is unclear.

Clones EP282 and EPMGHR2 are designated ‘research antibodies’, meaning they are not validated for clinical use, however several clinical laboratories reported utilising EP282 in diagnostic practice^31^ and studies assessing ROS1 status in breast cancer are increasingly using the EP282 clone.^40,41^ Indeed, these clones appear to demonstrate comparable performance to D4D6 and SP384 in the lung cancer literature.^31,42^ We found that clones EP282 and EPMGHR2 were positive in a substantial proportion of ILC (47% and 37%) and NST (49% and 74%) cases, respectively, with a range of staining intensities. Interestingly, EP282 and EPMGHR2 were positive in different patient cohorts, with EP282 being strongly associated with ER/PR negative, HER2 positive breast cancer, while EPMGHR2 was significantly associated with triple negative breast cancer. Furthermore, these two clones showed only moderate concordance, with observed agreement in 70% of ILC and 75% of NST cases, and AC1 values of 0.432 and 0.497 respectively. In clinical practice, this level of agreement is unacceptable for two assays designed to measure the same analyte, indicating a substantial level of unreliability and implying that at least one of the assays is nonspecific. Notably, EPMGHR2 binds within the tyrosine kinase (TK) domain of the ROS1 protein (Fig 1), which shares substantial homology with other TKs in the insulin receptor family, such as ALK.^43^ The target epitope of EP282 was not disclosed by the manufacturer.

Other studies also report markedly discordant results with clones D4D6 and EP282 in the breast cancer setting, while SP384 and EPMGHR2 have not been previously characterised in NST and ILC. Using D4D6, all 631 unselected breast cancer cases were ROS1 negative in one study.^44^ In contrast, two studies utilising the EP282 clone classed 33% (n=30)^41^ and 39% (n=137)^40^ of NST cases as ROS1 positive, which was associated with HER2 positivity in both studies. In contrast, a fourth study utilising a now-discontinued Santa Cruz antibody reported that 54% of NST cases (n=203) were ROS1 positive, which was associated with lower histologic grade, low proliferation index and ER positivity.^45^ Variability in study design, such as antibody clone selection, experimental conditions, antigen retrieval methodologies, detection systems and scoring methods, likely contribute to these heterogenous results, highlighting the importance of standardisation in biomarker testing and interpretation.

We suggest two potential explanations for the observed results. It may be that there is no significant upregulation of ROS1 protein expression in NST or ILC, as demonstrated by D4D6 and SP384, with both EP282 and EPMGHR2 showing nonspecific aberrant positivity in breast cancer. It is also possible that ROS1 is upregulated in breast cancer, however D4D6 and SP384 lack sensitivity for detecting ROS1 in this setting, while either EP282 or EPMGHR2 is appropriately sensitive (although both clones may still be nonspecific). To compare the ability of each antibody to accurately detect wild-type ROS1, we overexpressed ROS1 in two ROS1-negative cell lines. Our western blot data suggests that D4D6 can accurately detect full-length ROS1 in both cell lines, however, may also bind other proteins or variants, as evidenced by the presence of multiple bands in HEK293T cells. EP282 and EPMGHR2 were more specific in HEK293T cells, detecting a single band, but unsuccessful in HeLa, suggesting that ROS1 may undergo post-translational regulation influencing antibody binding and sensitivity. This data further highlights discrepancies across the antibody clones in accurately detecting ROS1 protein. Additional orthogonal validation approaches will be required to reconcile these disparate findings, to confirm antibody accuracy and understand the true status and biological significance of ROS1 protein expression in breast cancer.

This is the first study to conduct a comprehensive analysis of four ROS1 antibody clones across a large cohort of breast cancer cases with clinicopathological correlates. This study has several limitations. First, we can only report on the observed performance of the antibody clones, without drawing firm conclusions on analytical validity, which should be explored in future studies using orthogonal strategies. Second, TMAs were constructed from archival tissue where preanalytical variables could not be controlled, which may impact antibody clone performance. Third, analytical conditions were not constant across the four clones as immunohistochemistry was performed manually in-house for two clones (EP282 and EPMGHR2) and on automated staining platforms in diagnostic laboratories for the other two clones (D4D6 and SP384). Fourth, scoring was not blinded to antibody clone.

## Conclusions

In summary, we show that ROS1 antibody clones are not interchangeable in the breast cancer setting and highlight major discordances in ROS1 status according to antibody clone utilised. In addition, antibody clone selection influences the inferences made about the biological significance of ROS1 positivity in breast cancer. Confirmation of ROS1 antibody sensitivity and specificity is an important aim for future studies, prior to the assessment of the clinical validity of ROS1 as a predictive biomarker in breast cancer clinical trials.

## Data Availability

All data produced in the present study are available upon reasonable request to the authors

## Acknowledgements

We thank the patients and their families for their tissue donations and support. AS is the recipient of a Breast Cancer Trials Clinical Fellowship.

## Author Contribution

A.E.M.R, S.R.L and A.S conceptualised the study; A.E.M.R., S.R.L, P.T.S and A.S developed methodology and performed writing, review and revision of the paper; A.S, V.J, H.C, M.W, J.R.K, K.F, P.T.S., S.R.L., and A.E.M.R generated, analysed and interpreted data. All authors read and approved the final paper.

## Notes

Funding statement: AS is supported by Fellowship funding from Breast Cancer Trials.

### Competing Interest Statement

The authors have declared no competing interest.

### Funding Statement

Anna Sokolova received fellowship support from Breast Cancer Trials.

### Author Declarations

Ethics committees of The University of Queensland (2005000785) gave ethical approval for this work. Royal Brisbane and Womens Hospital (RBWH 2005/022) gave ethical approval for this work. UnitingCare Health (2006-39) gave ethical approval for this work. Public Health Act from the Queensland Government (CT_4352) gave approval for this study.

